# Characterizing clinical progression of COVID-19 among patients in Shenzhen, China: an observational cohort study

**DOI:** 10.1101/2020.04.22.20076190

**Authors:** Qifang Bi, Chengcheng Hong, Juan Meng, Zhenke Wu, Pengzheng Zhou, Chenfei Ye, Binbin Sun, Lauren M. Kucirka, Andrew S. Azman, Tong Wang, Jiancong Chen, Zhaoqin Wang, Lei Liu, Justin Lessler, Jessie K. Edwards, Ting Ma, Guoliang Zhang

## Abstract

**Background:** Understanding clinical progression of COVID-19 is a key public health priority that informs resource allocation during an emergency. We characterized clinical progression of COVID-19 and determined important predictors for faster clinical progression to key clinical events and longer use of medical resources.

**Methods and Findings:** The study is a single-center, observational study with prospectively collected data from all 420 patients diagnosed with COVID-19 and hospitalized in Shenzhen between January 11^th^ and March 10^th^, 2020 regardless of clinical severity. Using competing risk regressions according to the methods of Fine and Gray, we found that males had faster clinical progression than females in the older age group and the difference could not be explained by difference in baseline conditions or smoking history. We estimated the proportion of cases in each severity stage over 80 days following symptom onset using a nonparametric method built upon estimated cumulative incidence of key clinical events. Based on random survival forest models, we stratified cases into risk sets with very different clinical trajectories. Those who progressed to the severe stage (22%,93/420), developed acute respiratory distress syndrome (9%,39/420), and were admitted to the intensive care unit (5%,19/420) progressed on average 9.5 days (95%CI 8.7,10.3), 11.0 days (95%CI 9.7,12.3), and 10.5 days (95%CI 8.2,13.3), respectively, after symptom onset. We estimated that patients who were admitted to ICUs remained there for an average of 34.4 days (95%CI 24.1,43.2). The median length of hospital stay was 21.3 days (95%CI, 20.5,22.2) for cases who did not progress to the severe stage, but increased to 52.1 days (95%CI, 43.3,59.5) for those who required critical care.

**Conclusions:** Our analyses provide insights into clinical progression of cases starting early in the course of infection. Patient characteristics near symptom onset both with and without lab parameters have tremendous potential for predicting clinical progression and informing strategic response.

## Introduction

The epidemic of coronavirus SARS-CoV-2 has led to 1.6 million infections and over 100,000 deaths over 4 months after the first case was detected[1], causing severe shortage of essential medical supplies and equipment, medical staff, and hospital beds [2]. Complementary to data from COVID-19 epicenters like Wuhan (China) or Lombardy (Italy), data from places where healthcare capacity was not exceeded and patients were treated early and free of charge has the potential to shed light on the near complete clinical trajectory of cases. Clear characterization of COVID-19 clinical trajectory under the current standard of care informs planning for healthcare resource allocation during COVID-19 outbreaks and provides a basis that helps assess the effectiveness of new treatment and therapeutics.

Here, we use rich data on clinical progression of all cases diagnosed with COVID-19 diagnosed and treated in the only designated hospital in Shenzhen, China. Because all clinically confirmed cases, including a sizable portion detected through contact tracing, were required to be hospitalized for isolation purposes regardless of their clinical presentation and symptom profile, this dataset allows us to examine clinical progression of cases without the considerable selection bias typically seen in hospital-based studies. We estimate time from symptom onset to key clinical events, such as first clinical diagnosis, progression to severe clinical stages, development of acute respiratory distress syndrome (ARDS), admission to the critical care unit (ICU), and discharge. We also estimate duration hospitalized, in the ICU, and on ventilators. We determine the key predictors of faster clinical progression to a series of clinical events and longer use of healthcare resources.

## Methods

### Study design and study population

This single-centre, observational study was conducted at Shenzhen Third People’s Hospital, which is the designated hospital to treat all patients diagnosed with COVID-19 in Shenzhen. We prospectively collected data of all 420 patients diagnosed and hospitalized with COVID-19 in Shenzhen between January 11^th^ and March 10^th^, 2020, regardless of their clinical severity and symptom profile.

### Data collection

Epidemiological, demographic, symptom, laboratory, clinical, and outcome data were extracted from electronic medical records by multiple reviewers (BS, JC, JZ, PZ, JM, CH). All information was updated as of April 7^th^, 2020. Medical history was self-reported. Date of symptom onset before admission was self-reported and date of symptom onset after admission was recorded by an attending physician. We identified abnormality cutoffs for all laboratory results and recorded the date when such abnormalities were detected (See Table 1 for the complete list). We recorded the lowest cycle threshold values from the available RT-PCR testings and the date of testing. Smoking history was available for patients aged 60 years or older and was self-reported.

**Table 1.**
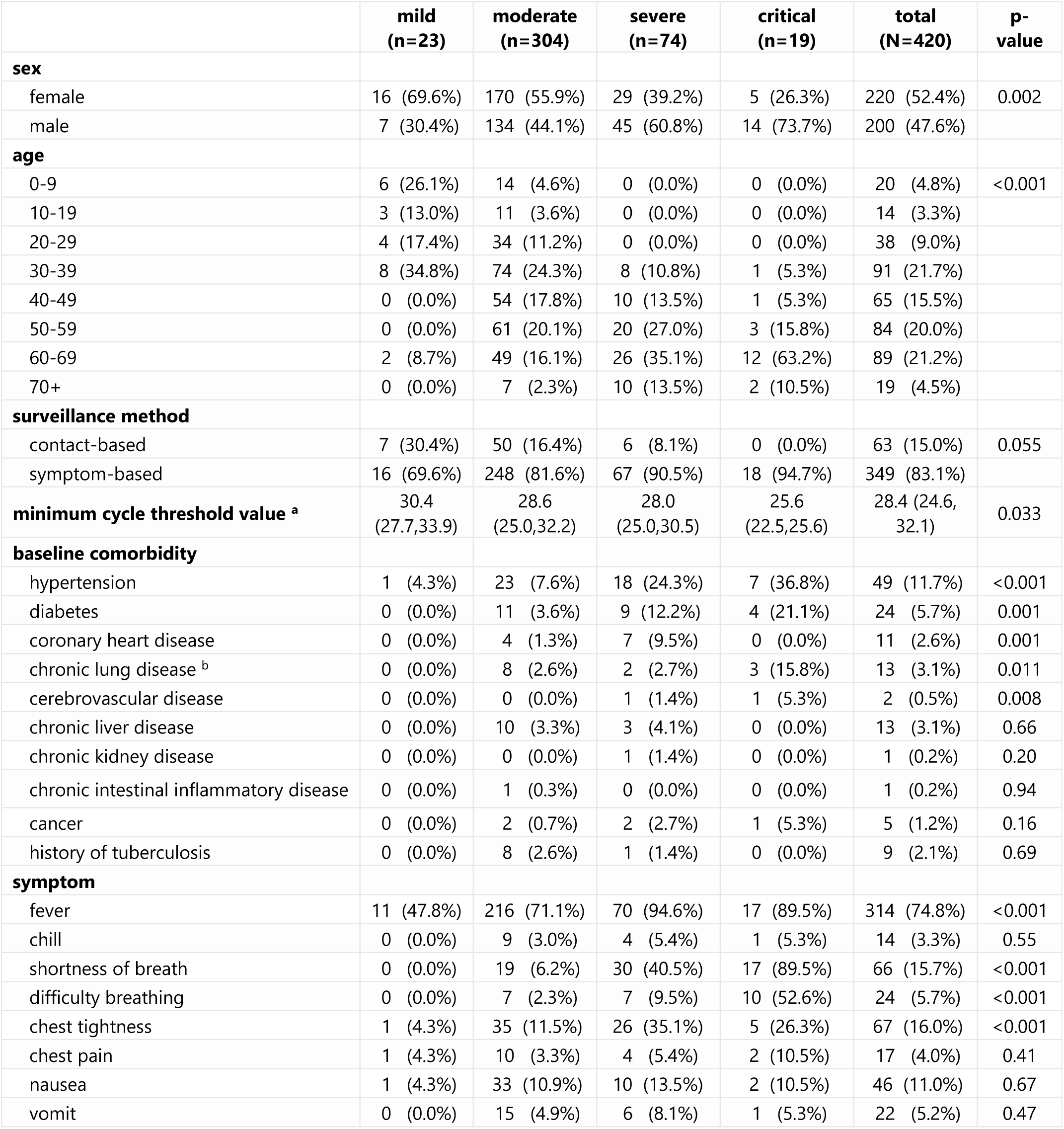
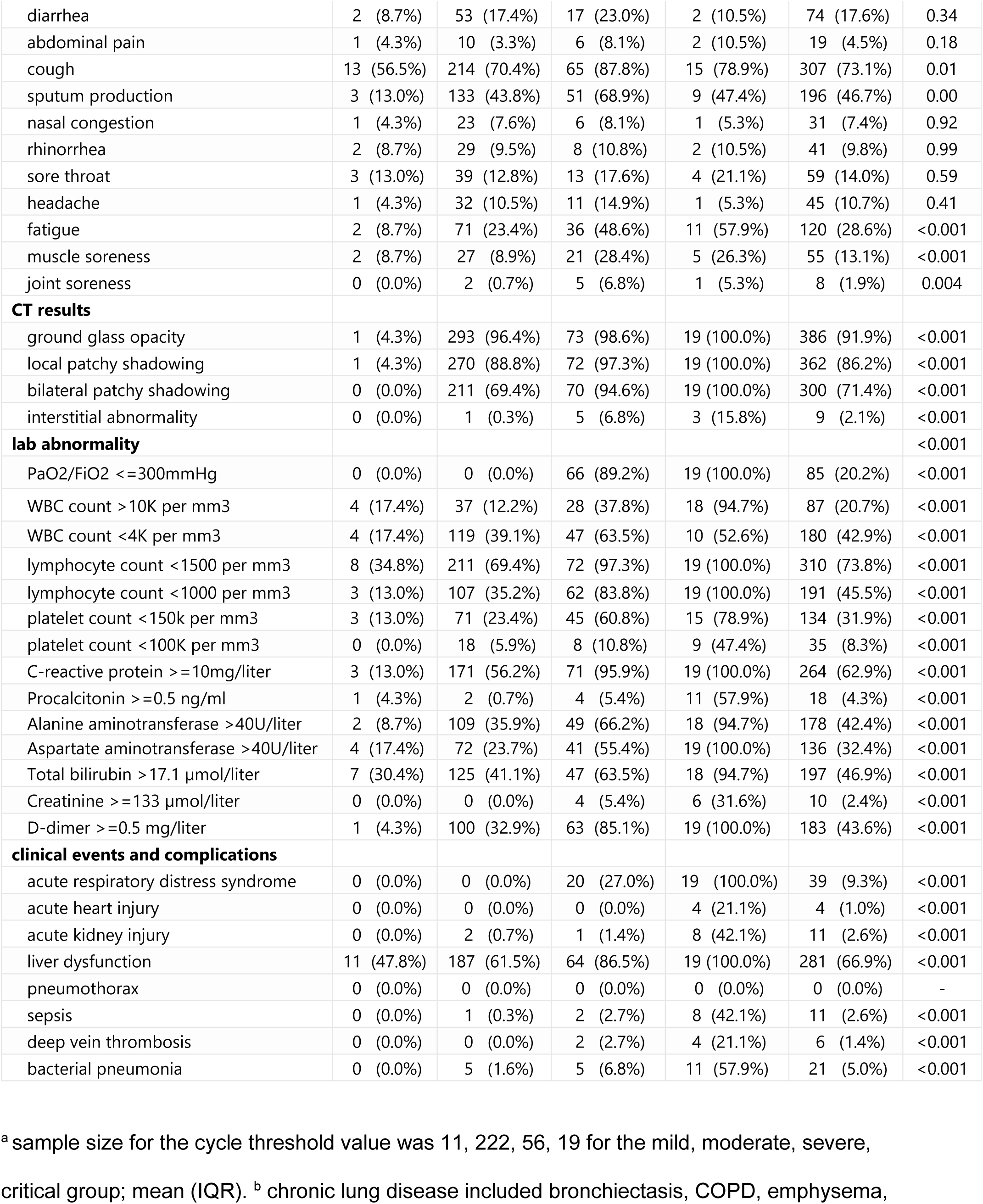
Demographic characteristics, surveillance method, baseline comorbidity, symptom profile, and lab and CT results by the highest clinical severity assessment. Comorbidities were self-reported on admission. P-values were assessed using χ^2^-tests

We recorded clinical severity at initial diagnosis and date of severity progression. Clinical severity was defined based on official guidelines and the definition was generally consistent over time (supplemental table 1 shows guidelines in each version) [3]. Patients were generally required to be hospitalized for 2 weeks for isolation purposes and were eligible for hospital discharge or transfer to a non-COVID ward for treatment if they met all four requirements: 1) no fever for over 3 days, 2) drastic improvement in respiratory symptoms, 3) pulmonary imaging showing significant reduction in inflammation, and 4) two consecutive negative RT-PCR results from respiratory sampling conducted over one day apart. [3]

The primary endpoints in this study include patients’ time from symptom onset to clinical progression beyond the moderate stage, ICU admission, invasive ventilator use, and hospital discharge. Secondary endpoints included time to when PaO_2_/FiO_2_ dropped under 300mmHg and time to developing ARDS.

### Statistical analyses

We estimated cumulative incidence of developing key clinical events in the presence of competing risks (i.e., death and hospital discharge) using the Aalen-Johansen estimator [4,5]. Outcomes included progression to severe stage, low PaO2/FiO2 ratio, ARDS, ICU admission, use of invasive ventilator, and hospital discharge.

Building upon the cumulative incidence of developing key clinical events, we used a non-parametric approach to estimate duration of hospitalization, duration in ICU among the patients admitted to the ICU, duration of invasive ventilator use among patients who require ventilator support (see Text S1 for detailed method). We used bootstrap simulation to construct confidence intervals. 95% confidence intervals were the 2.5th and 97.5th percentiles of the distribution of point estimates from the bootstrap samples.

We used competing risk regressions according to the methods of Fine and Gray [6] to estimate subdistribution hazard ratios comparing the rate of clinical progression between subgroups that were defined a priori (See Table 2 for the list of subgroups). We compared the rate of clinical progression to severe stage, ICU admission, ARDS, and hospital discharge between subgroups. Except for the models where time to hospital discharge was the outcome, hospital discharge and death were treated as competing events and end of study as administrative censoring for those still in treatment.

**Table 2:** The association of demographic characteristics, baseline comorbidity, initial symptoms, and initial lab results with rate of clinical progression to severe stage, acute respiratory distress syndrome, and ICU admission. We used competing risk regressions according to the methods of Fine and Gray to estimate subdistribution hazard ratios comparing the rate of clinical progression between subgroups.

|  | progression to severe stage |  |  | progression to acute respiratory distress syndrome |  |  | progression to ICU admission |  |  |
| --- | --- | --- | --- | --- | --- | --- | --- | --- | --- |
|  | sHR | 2.5% | 97.5% | sHR | 2.5% | 97.5% | sHR | 2.5% | 97.5% |
| **sex** |  |  |  |  |  |  |  |  |  |
| female | ref |  |  | ref |  |  | ref |  |  |
| male | 2.0 | 1.3 | 3.0 | 1.4 | 0.8 | 2.7 | 3.1 | 1.1 | 8.7 |
| **age** |  |  |  |  |  |  |  |  |  |
| 0-39 | ref |  |  | ref |  |  | ref |  |  |
| 40-59 | 4.5 | 2.1 | 9.3 | 6.2 | 1.4 | 28.1 | 4.4 | 0.5 | 39.6 |
| 60+ | 10.1 | 5.0 | 20.5 | 21.9 | 5.2 | 91.7 | 22.4 | 3.0 | 168.2 |
| **surveillance method** |  |  |  |  |  |  |  |  |  |
| contact-based | ref |  |  | ref |  |  | - | - | - |
| symptom-based | 2.7 | 1.2 | 6.2 | 3.4 | 0.8 | 13.9 | - | - | - |
| **cormorbidity** |  |  |  |  |  |  |  |  |  |
| hypertension | 3.2 | 2.1 | 5.1 | 3.6 | 1.8 | 6.9 | 4.6 | 1.8 | 11.6 |
| diabetes | 3.0 | 1.6 | 5.4 | 5.5 | 2.7 | 11.3 | 4.7 | 1.6 | 14.1 |
| coronary heart disease | 3.7 | 1.9 | 7.0 | 2.0 | 0.5 | 7.4 | 0.0 | 0.0 | 0.0 |
| chronic lung disease | 2.0 | 0.8 | 4.7 | 4.2 | 1.5 | 11.8 | 6.2 | 1.9 | 20.1 |
| cerebrovascular disease | 5.2 | 4.2 | 6.6 | 5.6 | 1.2 | 24.9 | 21.7 | 1.6 | 300.5 |
| **total comorbidity** |  |  |  |  |  |  |  |  |  |
| 0 | ref |  |  | ref |  |  | ref |  |  |
| 1 | 2.6 | 1.6 | 4.2 | 3.0 | 1.4 | 6.3 | 2.1 | 0.7 | 6.7 |
| 2,3 | 3.6 | 2.0 | 6.5 | 4.8 | 2.2 | 10.6 | 6.0 | 2.1 | 17.3 |
| early symptom |  |  |  |  |  |  |  |  |  |
| fever | 3.4 | 1.9 | 6.1 | 2.2 | 1.0 | 4.9 | 4.0 | 0.9 | 17.2 |
| cough | 2.0 | 1.2 | 3.1 | 1.5 | 0.8 | 3.0 | 1.5 | 0.6 | 3.8 |
| shortness of breath | 4.7 | 2.4 | 9.3 | 7.2 | 3.2 | 16.0 | 15.5 | 6.1 | 39.2 |
| muscle soreness | 3.2 | 2.0 | 5.2 | 3.0 | 1.5 | 6.1 | 2.2 | 0.8 | 6.7 |
| fatigue | 2.8 | 1.8 | 4.3 | 3.1 | 1.7 | 5.9 | 3.9 | 1.6 | 9.5 |
| lab results near symptom onset |  |  |  |  |  |  |  |  |  |
| PaO2/FiO2 <= 300mmHg | 38.3 | 14.6 | 100.8 | 16.7 | 7.4 | 37.8 | 22.3 | 8.4 | 58.8 |
| lymphocyte count < 1500 per mm3 | 2.1 | 1.4 | 3.1 | 2.1 | 1.1 | 4.0 | 3.6 | 1.3 | 10.0 |
| lymphocyte count < 1000 per mm3 | 2.1 | 1.4 | 3.3 | 2.9 | 1.5 | 5.5 | 4.5 | 1.8 | 11.0 |
| platelet count <150,000 per mm3 | 3.1 | 2.0 | 4.7 | 2.1 | 1.1 | 4.1 | 3.0 | 1.2 | 7.5 |
| C-reactive protein >= 10mg/L | 3.6 | 2.4 | 5.4 | 3.1 | 1.6 | 5.7 | 10.6 | 3.5 | 31.6 |
| D-dimer >= 0.5mg/L | 3.0 | 1.9 | 4.8 | 3.2 | 1.6 | 6.3 | 5.0 | 2.0 | 12.3 |

We used a flexible approach to stratify cases into three risk strata for the purpose of visualizing different clinical trajectories. We constructed a random survival forest model (RSF) and divided cases into low, medium, and high risk groups based on tertiles of RSF out-of-bag predictions (See Text S2 for methods). The candidate predictors used in the RSF model included 1) demographic information, 2) baseline comorbidities, 3) symptom profile, lab and CT results within 5 days of any symptom onset. We constructed another RSF model that stratified patients into risk groups without using lab or CT results near symptom onset. For both models, hospital discharge was treated as a competing event. All four deaths occurred after cases progressed beyond the severe stage, thus they were not treated as competing events. We calculated AUC over time since symptom onset (tAUC), providing a measure of model performance across all possible classification thresholds and based on the observed number of cases entering the severe stage by each time point [7].

## Results

### Characteristics of cases

Four-hundred and twenty cases were admitted and hospitalized at Shenzhen Third People’s Hospital between January 11th and March 10th, 2020 (Fig S1). On average, the first clinical diagnosis occurred 1.9 days (95% CI 1.6,2.3) and hospitalization occurred 4.2 days (95%CI 3.8,4.6) after symptom onset.

Of the 420 cases, there were approximately equal numbers of males (47.6%, n=200) and females (52.4%, n=220) (Table 1). Mean age of patients was 45 (IQR=34,60). Fifteen percent (63/420) of cases were detected through contact tracing, of which 9.5% (6/63) developed severe or critical disease. In contrast, 24.4% (85/349) of patients identified through symptom-based surveillance developed severe or critical disease. Patients identified through contact tracing were less likely to present fever or cough throughout the course of infection than patients identified through symptom-based surveillance (60%,38/63 contact-based vs. 19%,282/349 symptom-based patients never had fever; 38%,24/63 contact-based vs. 25%,87/349 symptom-based patients never had cough).

21.9% (92/420) of cases had at least one self-reported comorbidity on admission, with hypertension (n=49) and diabetes (n=24) being the most prevalent. Fever, cough, sputum production were the most common initial symptoms, with 68.6% (288) of patients showing fever, 60.0% (252) with cough, and 31.2% (131) with sputum production within 5 days of initial symptom onset (see Fig S2 for onset time distribution of each symptom).

At the initial clinical assessment, 23 patients (5.5%) were clinically mild, the vast majority (93.8%, n=394) were moderate, and only 3 patients were clinically severe or critical (Fig 1a).

**Figure 1:**
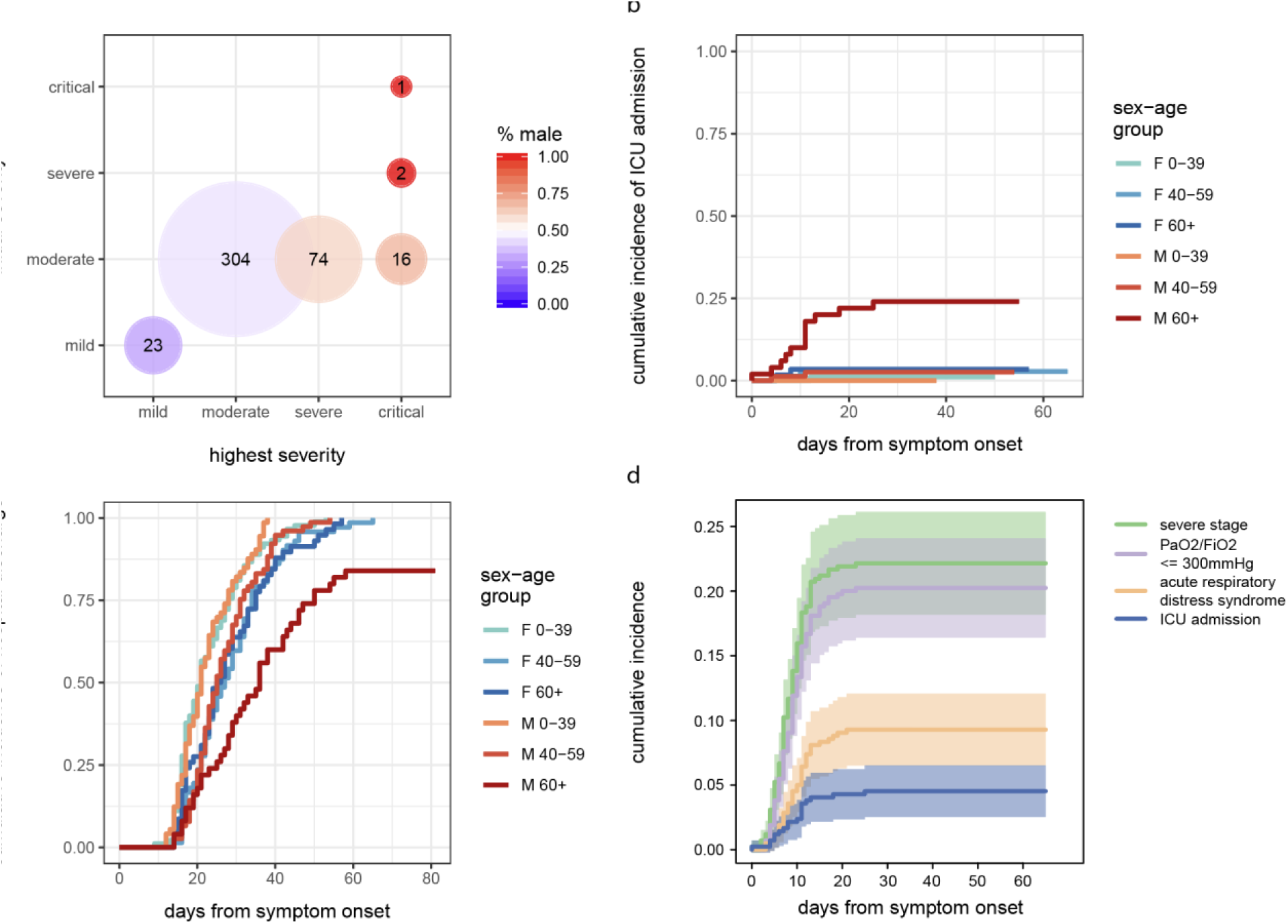
Cumulative incidence to clinical events and difference by age and sex. a) severity assessment at the initial diagnosis and the highest severity level during hospitalization of all 420 cases in Shenzhen. At the initial assessment, 93.8% (394/420) of cases had moderate clinical severity. Only 3 patients were either clinically severe or critical. 23 were mild. Of the 420 cases, 74 (17.6%) patients eventually became clinically severe and 19 (4.5%) eventually required ICU admission. All 23 mild cases during initial assessment stay in the mild category. b) cumulative incidence of ICU admission by age group and sex. c) cumulative incidence of hospital discharge by age group and sex. Clinical progression to ICU admission and hospital discharge was significantly different between males and females aged 60 and above. d) Cumulative incidence of advancing to severe stage, PaO2/FiO2 dropping below 300mmHg, requiring ICU admission, and developing acute respiratory distress syndrome.

### Clinical progression of cases

We estimated the proportion of the initially mild or moderate cases in each stage (mild/moderate, severe, ICU, death or discharge) over time following symptom onset, taking into account patients both transitioning into and out of each stage (Fig 2a, Text S3). The total number of patients in the severe stage reached its peak 12 days after symptom onset. Among the 417 patients who were classified as mild or moderate at the time of initial assessment, 21.6% (90/417) progressed to the severe stage. 9.6% (95%CI, 6.8%, 12.4%) progressed to the severe stage within 7 days after symptom onset, and 20.4% (95%CI, 16.5%, 24.3%) progressed within 14 days (Fig 1d and 2). Those who progressed to the severe stage progressed on average 9.5 days (95%CI 8.7,10.3) after symptom onset. Among the 417 patients who were classified as mild or moderate at the time of initial assessment, 8.6% (36/417) developed ARDS. 2.6% (95%CI 1.1%,4.2%) developed ARDS within 7 days from symptom onset, and 7.7% (95%CI 5.1%,10.2%) within 14 days. Those who developed ARDS developed ARDS on average 11.0 days (95%CI 9.7,12.3) after symptom onset.

**Figure 2:**
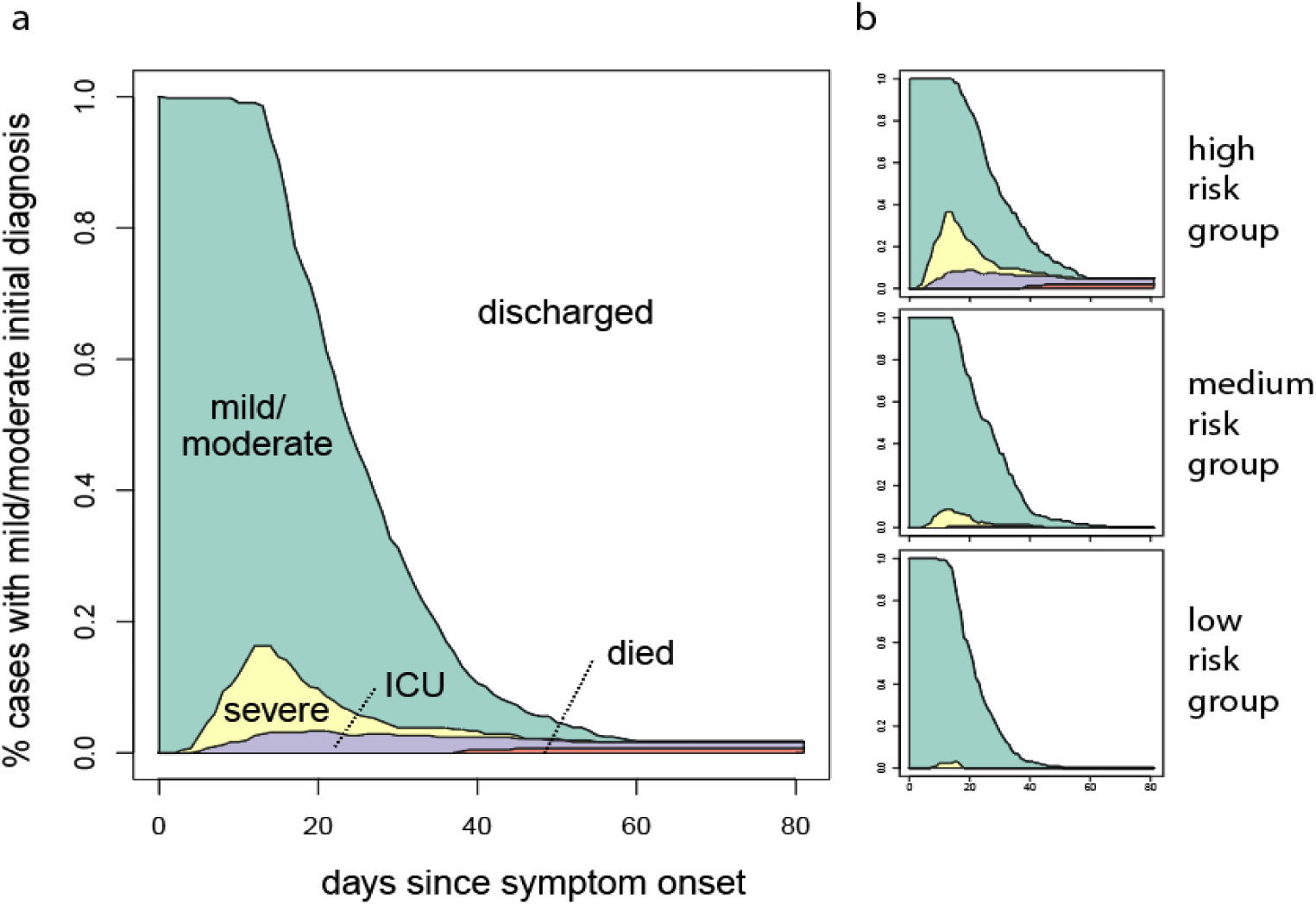
Clinical progression within 80 days following symptom onset for a) all cases with mild or moderate initial assessment, and b) cases in each of three risk subgroups obtained from random survival forest (RSF) a) From the top to the bottom, the four curves show the time-varying proportions of all admitted cases who 1) have not been discharged, i.e., still hospitalized or have died in hospital, 2) were severe, in the ICU, or have died, 3) were in the ICU or have died, and 4) have died. Successive differences between the four curves over 80 days from symptom onset were highlighted in distinct colors and show daily composition of cases in each of four stages (mild/moderate, severe, ICU, died). b) The candidate predictors used in the RSF model included 1) demographic information, 2) baseline comorbidities, 3) symptom profile, lab and CT results within 5 days of any symptom onset. The most important predictors of faster progression to the severe stage were low PaO2/FiO2 ratio, low platelet count, and high C-reactive protein concentration (Fig S7a; see Fig S7b for model performance). Risk stratification produced by an RSF model that excluded lab and CT results near symptom onset from the candidate predictors was highly similar to the risk stratification shown in b (Fig S6, Fig S7c-d).

As of April 7th, 19 patients had been admitted to ICU, among which 18 patients required invasive mechanical ventilation support, 4 patients died (1 patient died after initial hospital discharge with viral clearance), and 4 patients remained hospitalized in critical condition. We estimated that among the 417 patients who were classified as mild or moderate at the time of initial assessment, 3.3% (95%CI, 1.6%, 5.1%) of patients required ICU admission within 14 days from symptom onset (same for patients who required mechanical ventilators support). Those who required ICU admission were admitted into ICU on average 10.5 days (95%CI 8.2,13.3) after symptom onset. Using data from the 19 patients who were admitted into ICU, we estimated the average time in ICU was 34.4 days (95%CI 24.1,43.2) (Table S2). Using data from the 18 patients who required mechanical ventilator support, we estimated the average time on a ventilator was 28.5 days (95%CI 20.0,39.1).

The median length of hospital stay was 21.3 days (95%CI, 20.5, 22.2) for the mild or moderate cases who did not progress to the severe stage, and increased to 30.3 days (95%CI, 26.7,31.4) for cases who reached the severe stage but did not enter ICU and 52.1 days (95%CI, 43.3,59.5) for the cases who required ICU admission. Of note, patients in Shenzhen were required to be hospitalized for about 2 weeks for isolation. As a result, the duration of hospitalization for the mild or moderate cases was slightly inflated. However, patients with mild disease still have a long clinical course, because the majority (79%, 331/417) of the cases that were initially mild or moderate were hospitalized for more than 14 days (66%, 277/417 for more than 16 days). All patients who were clinically mild at the time of initial assessment stayed mild until discharge (Fig 1a).

### Characteristics associated with difference in clinical progression

We then identified a priori-defined patients’ characteristics that were associated with faster clinical progression. We found that having hypertension and diabetes at baseline was strongly associated with faster clinical progression to various clinical events, including progression to the severe stage (sHR=3.2, 95%CI 2.1,5.1 for hypertension and sHR=3.0, 95%CI 1.6,5.4 for diabetes), to developing ARDS (sHR=3.6, 95%CI 1.8,6.9 for hypertension and sHR=5.5, 95%CI 2.7,11.3 for diabetes), and to ICU admission (sHR=4.6, 95%CI 1.8,11.6 for hypertension and sHR=4.7, 95%CI 1.6,14.1 for diabetes) (Table 2). Having more baseline comorbidities was also associated with a higher rate of clinical progression to these events (Table 2). Although many lab abnormalities measured within 5 days of symptom onset were strongly predictive of faster clinical progression, including low lymphocyte count, low platelet count, high concentration of C-reactive protein, and high concentration of D-dimer, notably, a low PaO2/FiO2 ratio close to symptom onset was very strongly associated with faster clinical progression. We observed a 22.3 times (95%CI 8.4, 58.8) increase in the subdistribution hazard of ICU admission among those with early measures of low PaO2/FiO2 ratio (Table 2).

Older age was one of the most important predictors of faster clinical progression (Table 2). All four patients who died were male over the age of 60. About half (46.3%, 50/108) of cases aged 60 or above progressed to the severe or critical stage (Table 1). Although the vast majority of those under the age of 40 did not progress beyond the moderate stage, 6% (9/163) of cases in this younger age group became clinically severe or critical and none of them had any known underlying comorbidities.

Difference in clinical progression by sex was mostly driven by the difference among older patients. We observed a notable difference in progression to require ICU admission between males and females aged 60 or above; males had a 10-fold increase in the subdistribution hazard of ICU admission compared to females in the same older age group (sHR=10.5, 95%CI 1.0, 108.6), despite the similar sex-specific age distribution in this age group (males: mean 67, IQR 63,69 vs. females: mean 65, IQR 62,66) (Table 1, Fig 1b). Differences in baseline comorbidities between older males and females did not explain the disparity; after adjusting for having any underlying comorbidity, the subdistribution hazard ratio remained unchanged (Table S3; Fig S5). Adjusting for smoking history did not explain the disparity by sex among those aged 60 or above. We did not observe a significant difference in time to ICU admission between males and females under the age of 60 (sHR=0.7, 95%CI 0.1,4.3). Similarly, we found that males aged 60 or above had a lower rate of hospital discharge compared to females in the same age group, and no significant difference by sex in the younger age group (Fig 1c). However, we did not find significant disparities in clinical progression to severe stage or to developing ARDS by sex in any age group (Table S3).

Using linear regression, we found that the minimum RT-PCR cycle threshold values for the severe cases were significantly lower than the mild cases after adjusting for time of sample collection from symptom onset. We observed a general trend of lower minimum cycle threshold values in patients with more severe clinical presentation and in patients in the older age group though the association with age was not statistically significant (Fig S4).

### Risk strata for clinical progression

Based on the random survival forest results, the most important predictors of faster progression to the severe stage were low PaO2/FiO2 ratio, low platelet count, and high C-reactive protein concentration (Fig S7a). We observed very different clinical trajectories of patients in each risk group (Fig 2b), highlighting the effectiveness of risk stratification produced by the RSF model. In the low-risk group, no case required ICU admission and only 3% (95%CI, 0.1%,6%) of cases became severe within 14 days from symptom onset. Whereas in the high-risk group, we estimated that 43% (95%CI, 35,52) cases became severe and 9% (95%CI, 4%,14%) required ICU admission within 14 days from symptom onset. All but one case that required ICU admission were classified into the high-risk group, and we estimated that the duration in ICU for those in the high-risk group was 35.1 days (95%CI, 26.1,45.1) (Table S2). Risk stratification produced by an RSF model that excluded lab and CT results from the candidate predictors was highly similar to the risk stratification produced by the model that considered lab and CT results, and it also differentiated the clinical trajectory of patients well (Fig S6, Fig S7c-d).

## Conclusions

The analysis of clinical data from patients diagnosed with COVID-19 in Shenzhen Third People’s hospital provides insights into clinical progression of cases starting early in the course of infection. We estimate the proportion of cases in each severity stage over 80 days following symptom onset. The overall trajectory of clinical progression of cases in Shenzhen were similar to estimates from COVID-19 epicenters, with mean time to ICU admission and to developing ARDS to be around 10 days [8,9,10]. We show that COVID-19 has a long clinical course even for those with mild disease (~20days), and time to recovery is close to 2 months for those who required ICU admission (~52days). Patients detected through contact tracing had milder diseases at initial assessment and were less likely to progress to the severe stage than those detected through symptom-based surveillance. Patient characteristics previously reported to be associated with ARDS and death [11] including hypertension, diabetes, and various lab indicators were also highly predictive of faster clinical progression.

Previous studies have reported substantial differences in risk of developing severe disease by sex and age [8][12][13]. We further show faster clinical progression among males than females that were primarily driven by the striking difference in the older age group independent of differences in baseline comorbidities or smoking history. Immunopathogenesis has been proposed to play a role in predisposing certain subgroups to be more at risk to severe disease than others. Understanding the underlying mechanism will be key to developing a safe vaccine and designing a safe vaccination strategy [14].

This study has several limitations. Dates of symptom onset were extracted from physicians’ notes that were not recorded to explicitly ascertain information on symptom onset. When the date of symptom onset could not be determined, we assumed onset date was the date of initial diagnosis. We performed sensitivity analyses to identify patient characteristics close to initial diagnosis that are predictive of clinical progression to severe stage and ICU admission and the results remained qualitatively the same (Table S5). We also performed the RSF analyses using symptom profile and lab results within 5 days of initial diagnosis as candidate predictors, and the integrated AUC remained relatively unchanged (Fig S8). Our estimates of duration of ICU stay and ventilator use were somewhat imprecise, likely due to the small sample size for those reaching the critical stage. Even though estimating duration using our non-parametric approach showed improvement in precision compared to times estimated using parametric accelerated failure time models, the estimated time within subgroups needs to be interpreted with caution because of the small sample size (Table S4). Finally, the association presented between clinical progression and patient characteristics should not be interpreted as causal given the variation in treatment and numerous confounders that were not accounted for in this study.

We demonstrate that patient characteristics near symptom onset have tremendous potential to inform COVID-19 triage, grouping patients into risk sets with different outlook of clinical progression. While our RSF model performs well based on out-of-bag predictions, we would be highly cautious of triaging patients in other settings using the important variables identified here due to our limited sample size. However, this is an important first step towards an applicable triage risk screening tool once well recorded data on clinical courses for more patients become available. Strategic response and allocation of medical resources for ongoing outbreaks may also benefit from a dynamic risk scoring system that incorporates new patient-level lab and symptom information as it is updated over time.

In conclusion, we provided quantitative characterization of the clinical progression of COVID-19 beginning from early clinical stages. Our estimates form the basis for assessing effectiveness of new treatments and inform planning for healthcare resource allocation during COVID-19 outbreaks.

## Data Availability

The data that support the findings of the study are not publicly available. Summary data may be available from the authors upon reasonable request with permission from Shenzhen Third People's Hospital.

## Acknowledgements

We thank all patients and their families involved in the study; as well as the front line medical staff and public health workers who collected this critical data. GZ was funded by the National Science and Technology Major Project for Control and Prevention of Major Infectious Diseases of China (No. 2017ZX10103004). CH, PZ, CY, BS, TW, JC, TM were funded by the Emergency Response Program of Harbin Institute of Technology (HITERP010) and Emergency Response Program of Peng Cheng Laboratory (PCLERP001). JL and QB were funded by a grant from the US Centers for Disease Control and Prevention (NU2GGH002000).

## Author Contributions

QB, Z Wu, JE conducted statistical analyses and drafted the figures

QB, Z Wu, JE, AA, JL, LK, CH, JM, GZ drafted and provided critical feedback on the manuscript

BS, JC, JZ, PZ, CH, JM, Z Wang, CY extracted the data

QB, TM, JL, LL, GZ, TW conceived the study

## Ethics Statement

This work was conducted in support of an ongoing public health response, hence was determined not to be human subjects research after consultation with the Johns Hopkins Bloomberg School of Public Health IRB. The study was approved by the ethics committees of Shenzhen Third People’s Hospital.

